# PERSONALIZED GLYCEMIC RESPONSES TO FOOD AMONG INDIVIDUALS WITH TYPE 2 DIABETES IN INDIA: DEVELOPMENT OF A MACHINE LEARNING PREDICTION MODEL

**DOI:** 10.1101/2024.10.20.24315560

**Authors:** Niteesh K. Choudhry, Shweta Priyadarshini, Jaganath Swamy, Mridul Mehta

## Abstract

**Objective:** Emerging global evidence demonstrates marked inter-individual differences in post-prandial glucose response (PPGR) although no such data exists in India and prior studies have primarily evaluated PPGR variation in individuals without diabetes. This study sought to develop a machine learning model to predict individual PPGR responses to facilitate the prescription of personalized diets for individuals with type 2 diabetes.

**Research Design and Methods:** Adults with type 2 diabetes and a hemoglobin A1c (HbA1c) ≥7 were enrolled from 14 sites around India. Subjects wore a continuous glucose monitor and logged meals. PPGR was calculated for each meal, based on the incremental area under the curve, and a machine learning predictor of PPGR was developed using stochastic gradient boosting regression. Model calibration and discrimination was assessed using a Pearson product moment correlation and area under a receiver operating curve (AUC), respectively, and its performance was compared to models based only on meal carbohydrate and calorie content.

**Results:** The study included data from 488 patients (mean age 52.5 years, 36% female, mean duration of diabetes 6.4 years, mean hemoglobin A1c 8.16%). Mean PPGR to common foods varied substantially (e.g. PPGR for “aloo paratha with curd” ranged from 10 to 170 mg/dl*h). PPGR values predicted by the machine learning model were highly correlated with observed PPGR (r=0.69) and model calibration was substantially stronger than for a model based only on calorie (r=0.57) or carbohydrate (r=0.39) content. The machine learning model also demonstrated very strong discriminative ability (AUC 0.80).

**Conclusions:** A machine learning model built with nutritional content, health habits, biometric information and common laboratory data produced highly accurate individualized predictions of PPGRs that substantially outperformed predictions based upon calorie and carbohydrate content. These results could be used to facilitate the delivery of personalized medical nutritional therapy as is widely recommended by type 2 diabetes practice guideline in India and globally.

---

As of 2023, more than 100 million Indians have diabetes, representing more than 11% of the population.(1) An additional 136 million Indians have prediabetes.(1) These numbers are anticipated to continue to grow rapidly.(2) The rising prevalence of this condition in India is believed to be the result of changing diets, increasingly sedentary occupations, lower levels of physical activity in the context of urbanization, and rapidly increasing rates of obesity.(3)

Along with medications and physical activity, every major clinical practice guideline globally emphasize diet as a key tenant of effective blood sugar control.(4–7) These recommendations are particularly relevant in the Indian context given the centrality of carbohydrates,(8) in particular white rice and refined wheats,(9) in the Indian diet. Like in other jurisdictions, Indian guidelines call for the individualization of meal planning, which is sometimes referred to as “Medical Nutritional Therapy”.(7) Personalization of dietary plans are generally based upon broad constructs like age, activity level, health status and preferences, and, for all patients, tend to emphasize calorie reduction and minimization of refined carbohydrate(10) in favor of the consumption of non-starchy vegetables and foods that are high in protein.(11) Nutritional recommendations have, thus far, not incorporated more personalized approaches to reflect substantial inter-individual differences in post-prandial glucose response (PPGR) including to foods that are relatively high in carbohydrate or calories.(12–16)

The existing literature on PPGR variability comes mostly from individuals without type 2 diabetes.(17) Further, there have been no large-scale studies characterizing food responsiveness among Indians. Given the unique features of diabetes among South Asians, which often referred to as the “South Asian Phenotype”,(18–20) there are very likely to be differences in PPGR in India as well, just as have been observed in Indians’ responses to some diabetes medications.(21) Accordingly, we aimed to develop a machine learning model based upon nutritional content, patient-reported health habits, biometric data and commonly-available laboratory data to predict individual PPGR responses with the ultimate goal of facilitating the prescription of personalized diets for individuals with type 2 diabetes.

## METHODS

We conducted a prospective cohort study to evaluate PPGR among individuals with type 2 diabetes in India. This study was approved by the ethics committees at all institutions enrolling patients and is registered with Clinical Trials Registry-India (CTRI/2022/02/040619). All subjects provided written informed consent. The authors are responsible for the design, conduct and analysis of this study and all met International Committee of Medical Journal Editors (ICMJE) criteria.

The study protocol was published previously.(22) Study enrollment began May 3, 2022, and was terminated on June 19, 2024 because of slower than anticipated enrollment and funding limitations.

### Study setting

This trial was conducted at 14 outpatient clinics in geographically distinct regions across India. Sites were included if they specialized in the care of individuals with diabetes, had an established research infrastructure for the conduct of diabetes-related studies, and a sufficient volume of potentially-eligible patients.

### Eligible subjects and enrollment

We included individuals age ≥ 18 and <75 years with physician-diagnosed type 2 diabetes receiving treatment ≥1 oral hypoglycemic agents, who had a hemoglobin A1c (HbA1c) value ≥ 7.0% measured within the 30 days prior to enrollment. Subjects were required to have a mobile phone capable of running the protocol-specified applications and to have functional English literacy. Patients were excluded if they were receiving prandial insulin including a continuous insulin infusion pump, were pregnant or planning to become pregnant, had an estimated life expectancy ≤ 12 months, had active cancer, had a myocardial infarction or stroke in the prior 6 months, were receiving or had planned to initiate dialysis for end-stage renal disease, were using oral or intravenous steroids or had any contraindication to using a continuous glucose monitor (CGM). Concurrent treatment with an injectable non-insulin agent was permitted.

Potentially eligible patients were identified from clinic records and underwent in-person screening to confirm eligibility and obtain written informed consent. As previously described,(22) consenting patients provided sociodemographic information and medical history, completed baseline surveys,(23–27) had biometric data collected including vital signs and body measurements and had blood samples including a complete blood count, HbA1c, blood electrolytes, creatinine, cholesterol, and urinalysis.

After completing baseline assessments, subjects were fitted with an Abbott Freestyle Libre CGM sensor, were provided with a Xiaomi Mi Band Smart Wristband heart rate monitor and a Roche Accu-Chek glucometer with testing supplies, and dietary supplements to be consumed with their standardized meals (see **Follow-up procedures** below). Study-specific applications were downloaded on to the subjects’ smartphones to allow them to log dietary intake and synchronize their CGM and heart rate monitors. As a back-up, subjects were given a paper dietary logbook and a Freestyle Libre CGM reader with which to collect protocol-specified data.

### Follow-up procedures

Subjects were followed for 14 days during which they logged their full dietary intake including all standardized test meals and free-living foods (including snacks), beverages (including water) and medications. Participants also logged all exercise.

Subjects were required to consume protocol-specified meals consisting of vegetarian breakfast foods that varied in their proportion of carbohydrate, fiber, protein, and fat. On other days, subjects were asked to consume normal foods with protocol-specified constraints. For example, on different days, subjects varied the types of mixed protein (e.g., different types of lentils with or without added protein), the order in which foods were consumed (e.g., protein before carbohydrate v. protein with carbohydrate), drank water before their meal, walked after eating or ate what they perceive to be a healthy meal. Where applicable, subjects were given several options as to which of their usual foods were acceptable for each protocol specified food modification. Details about the dietary protocol have been previously published.(22)

### Primary outcome

The study’s primary outcome was PPGR which was calculated for each logged meal by estimating the incremental area under the curve (iAUC) from continuous glucose measurements following the method of Wolever and Jenkins,(28) as adapted by Zeevi et al,(13) Mendes-Soares et al(15) and Berry et al.(29) Prior to conducting analyses, meals logged less than 30 minutes apart were merged and meals logged within 90 min of other meals removed. Consistent with prior PPGR prediction studies,(15) meals that were very small (<15 g and <70 calories) or very large (>1 kg) or which had implausibly low PPGR values (i.e. a PPGR < 5 mg/dl*h after consuming ≥ 40g of carbohydrates) were excluded as they likely represent inaccurate food logging. To reduce noise, the median of all glucose values from the 30-minute period prior to the meal was taken as the initial glucose level, above which the incremental area was calculated. Meals that had incomplete glucose measurements in the time window of 30 minutes before and 2 hours after the logged mealtime were filtered out.

### Statistical analysis plan

Descriptive statistics were used to describe the characteristics of the study population, to plot the range of carbohydrates consumed, calories consumed and PPGRs observed as well as to estimate the Pearson product moment correlation between PPGR and the nutritional composition of the logged meals.

A machine learning predictor was developed based on stochastic gradient boosting regression (XGBoost, version 2.2.1)(30) using the XGBRegressor class. PPGR was predicted as the sum of predictions from thousands of decision trees. Trees were inferred sequentially, with each trained on the residual of all previous trees. While we collected a wide variety of data from study participants, including meal content, demographics, health habits, baseline laboratory values, as well as CGM, heart rate and activity data, we were specifically interested in developing a model that relied on nutritional content, self-reported health habits (for example, exercise and sleep patterns), biometric information (for example, weight and waist size), and laboratory data commonly available to patients with type 2 diabetes in India (for example, HbA1c).

Model performance was assessed with measures of calibration and discrimination.(31) Model calibration was assessed by plotting the predicted and observed PPGR values. We then held out 30% of the sample and used 5-fold cross-validation on the remaining sample. In this approach, the cross-validation participants were divided into 5 groups and the model was trained on the other 4 groups. Random data sets of the same size as the original were then sampled with replacement from the original cross-validation data set, and the entire training and validation process was repeated. Prediction results were aggregated and then the accuracy of the resultant model was assessed in the held out 30% of subjects by calculating a Pearson product moment correlation between the predicted and measured PPGRs. The standard error for the calculated performance was assessed using at least a thousand iterations of bootstrapping until the errors stabilized.

Following the methods of Mendes-Soares et al,(15) discrimination was assessed using a binary cut-point for PPGR, set at the 50^th^ percentile of all observed PPGR values, plotting a receiver operative curve (ROC) and then calculating the area under the ROC (AUC), or equivalently a c-statistic, which evaluates whether the model was more likely to predict a high PPGR among those foods where a high PPGR was actually observed than for those foods with a low observed PPGR. For comparison, the AUC for prediction models based only on the carbohydrate and calorie content of the meal, consistent with standard dietary approaches used by patients with type 2 diabetes, was also calculated.(4)

### Sample size considerations

We planned to collect data from 1000 individuals and estimated that the collected data from them would provide 80% power to detect correlations of a magnitude of r=0.13 (R^2^=0.017) with p<0.005. Because of slower than anticipated enrollment, funding limitations required us to terminate by June 19, 2024, by which time we had recruited 512 patients. With the data from these individuals we estimated that we would still have 80% power to detect correlations of a magnitude of r=0.17 (R^2^=0.029) with an p<0.005.

## RESULTS

### Characteristics of study participants and data collected

We recruited 512 patients of whom 488 (95.3%) had evaluable data to be included in the current study. Study participants had a mean age of 52.5 years, 36% were female, they been diagnosed with type 2 diabetes for a mean of 6.4 years and had a baseline HbA1c of 8.6%. Other characteristics of study participants are shown in **Table 1**.

**TABLE 1:** Baseline characteristics of study participants.

| **VARIABLE** | **VALUE** |
| --- | --- |
| **Sociodemographic Characteristics** |  |
| Age, mean (SD) years | 52.5 (1) |
| Female sex, % | 36% |
| College or higher education, % | 44.5% |
| Diet, % |  |
| Vegan | 5.4% |
| Vegetarian | 33.9% |
| Pescatarian | 8.8% |
| Partial non-vegetarian | 7.9% |
| Non-vegetarian | 44.0% |
| **Diabetes Characteristics** |  |
| Duration of diabetes, mean (SD) years | 6.4 (0.6) |
| Diabetes treatments |  |
| Oral hypoglycemics only | 95.5% |
| Oral hypoglycemics + non-insulin injection | 0.2% |
| Oral hypoglycemics + basal Insulin | 4.3% |
| **Biometric Measurements** |  |
| Body mass index, mean (SD) kg/m<sup>2</sup> | 26.9 (0.51) |
| Waist to hip ratio, mean (SD) | 0.99 (0.04) |
| Systolic blood pressure, mean (SD) mmHg | 126.0 (1.1) |
| **Laboratory values** |  |
| Hemoglobin A1c, mean (SD) % | 8.6 (0.2) |
| Cholesterol, mean (SD) mg/dL |  |
| Total | 170 (4.1) |
| Low-density lipoprotein | 128 (4.0) |
| Triglycerides | 213 (16.9) |
| Creatinine, mean (SD) mmol/L | 0.85 (0.04) |

Subjects logged 22,967 meals, consisting of 9.2 million kilocalories, and provided 543,442 CGM readings. The mean number of calories and carbohydrates per meal were 400.5kcal and 58.8g, respectively. The distribution of calories consumed, carbohydrates consumed and PPGR values are shown in **Figure 1**.

**FIGURE 1:**
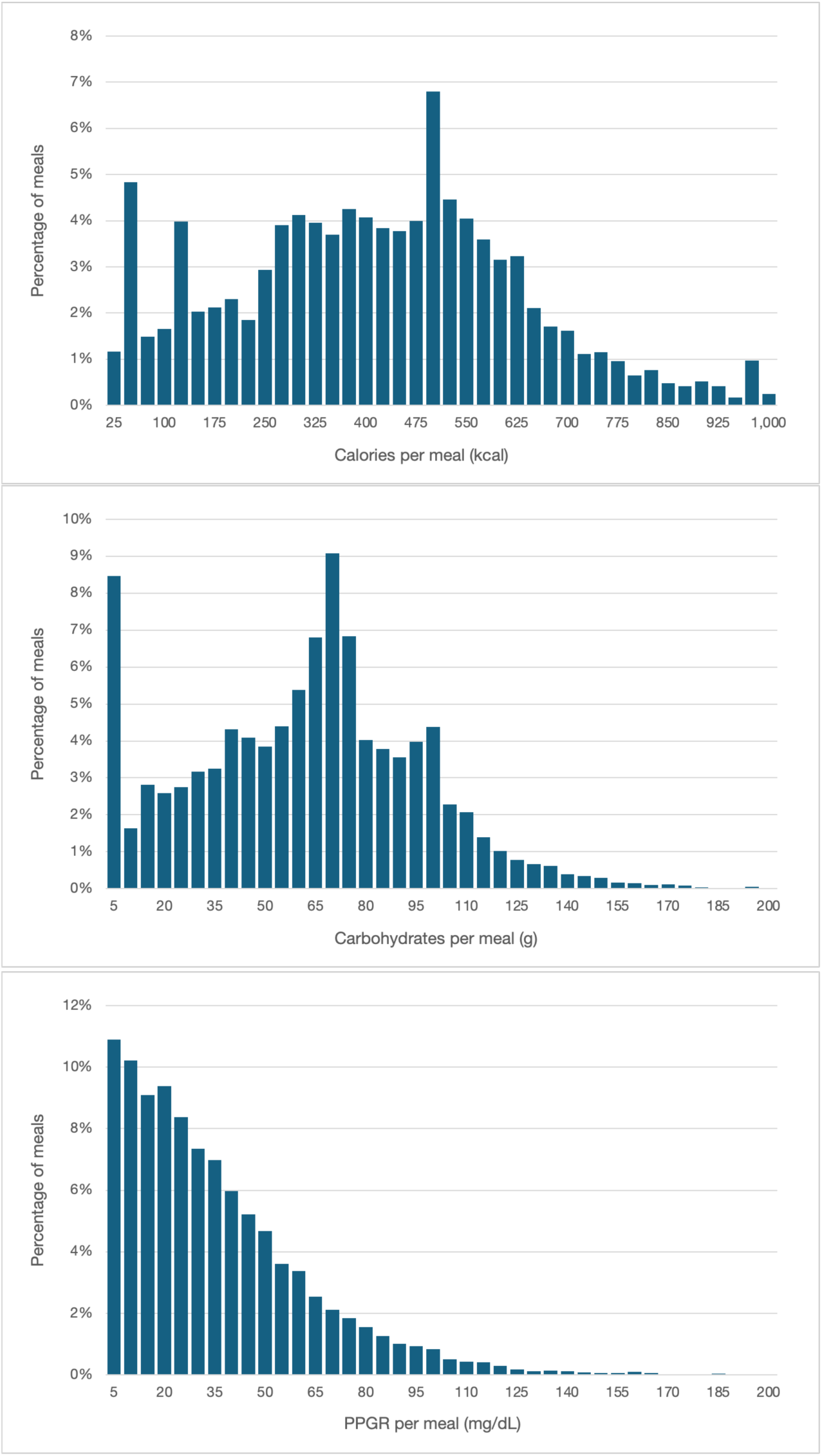
Distribution of carbohydrates consumed, calories consumed and PPGR among logged meals. The plots display the range of calories (top panel), carbohydrates (middle panel) and PPGR (bottom panel) per meal among study participants.

### Relationship of PPGR to nutritional composition of foods

The relationship between PPGR and nutritional content of foods consumed by study participants is shown in **Figure 2**. Mean PPGR increased with carbohydrate content (**Figure 2 top panel**), defined as the percentage of meal calories coming from carbohydrates, but the overall correlation was weak (r=0.25, p<0.001). The ratio of meal carbohydrates to protein had a weak negative correlation with PPGR (r=-0.05, p<0.001; **Figure 2 middle panel**) and PPGR was uncorrelated with the ratio of meal carbohydrate to fat (r=0.00, p<0.89 **Figure 2 bottom panel**).

**FIGURE 2:**
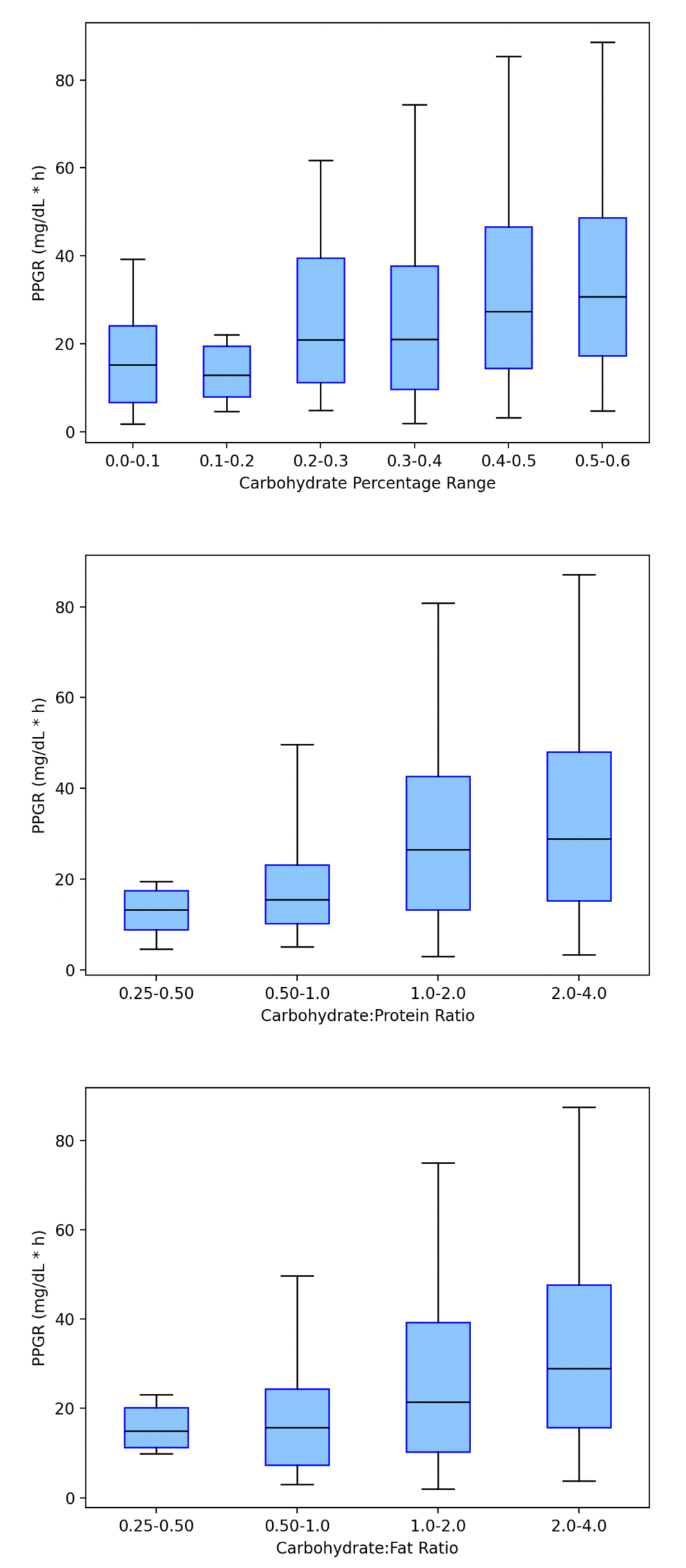
Relationship between PPGR and food components of logged foods among study participants. The plots show the relationship between PPGR and carbohydrate content (top panel), carbohydrate to protein ratio (middle panel) and carbohydrate to fat content (bottom panel) Each boxes show the 25^th^ percentile (bottom of box), median (box midline), and 75th percentile (top of box). The error bars represent the 5^th^ to 95^th^ percentile range.

As with responses to carbohydrates and other macronutrients described above, we observed substantial variation in PPGR to whole meals (**Figure 3**). **Figure 3 top panel** shows the mean PPGR responses to three common Indian foods consumed by study participants (masala dosa: PPGR range 10 to 150 mg/dl*h, aloo paratha with curd: PPGR range 10 to 170 mg/dl*h, and a vegetable sandwich with milk: range PPGR 14 to 121 mg/dl*h,). **Figure 3 bottom panel** shows PPGRs for 4 illustrative subjects who ate aloo paratha with curd, for which a standard serving contains 492 kcal, 68 grams of carbohydrates, 14 grams of protein and 21 grams of fat. The observed PPGRs for these 4 subjects were 24 mg/dl*h, 41 mg/dl*h, 65 mg/dl*h and 89 mg/dl*h.

**FIGURE 3:**
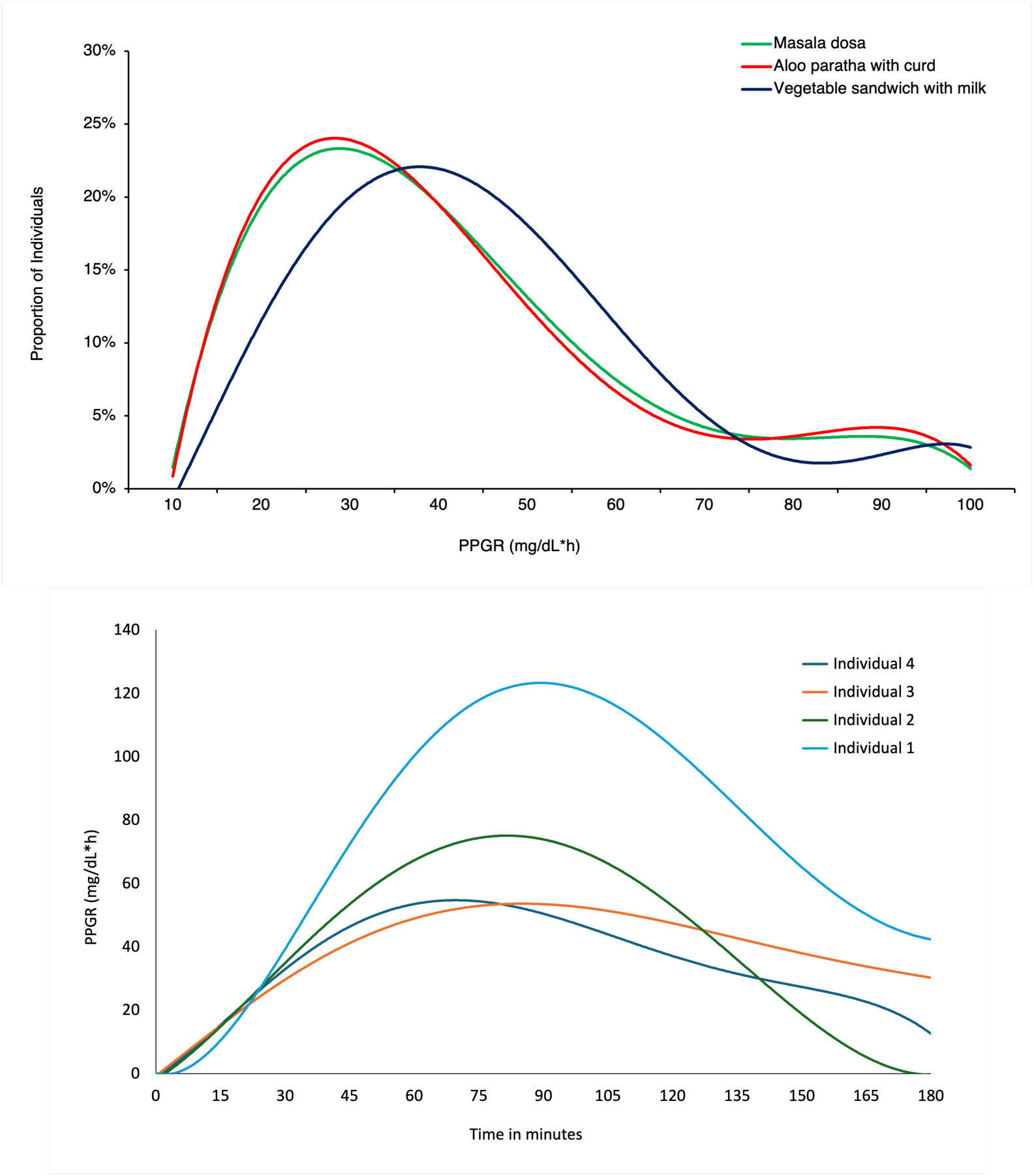
Variability in PPGR to common Indian foods among study participants. The plots show the distribution of PPGRs observed for 3 common foods among all study participants (top panel) and PPGRs for 4 illustrative patients in response to aloo paratha with curd (bottom panel). PPGRs values in the top panel were winsorized at 100 mg/dl*h for graphical representation.

### Performance of machine learning based PPGR prediction model

The relationship between observed PPGRs and those predicted by the machine learning model are shown in **Figure 4 top panel**, along with models based only on calorie content (**Figure 4 middle panel**) and carbohydrate content (**Figure 4 bottom panel**). PPGR values predicted by the machine learning model were highly correlated with observed PPGR (r=0.69, p<0.001) and model calibration was substantially stronger than for a model based only upon calorie (r=0.57, p<0.001) or carbohydrate (r=0.39, p<0.001) content.

**FIGURE 4:**
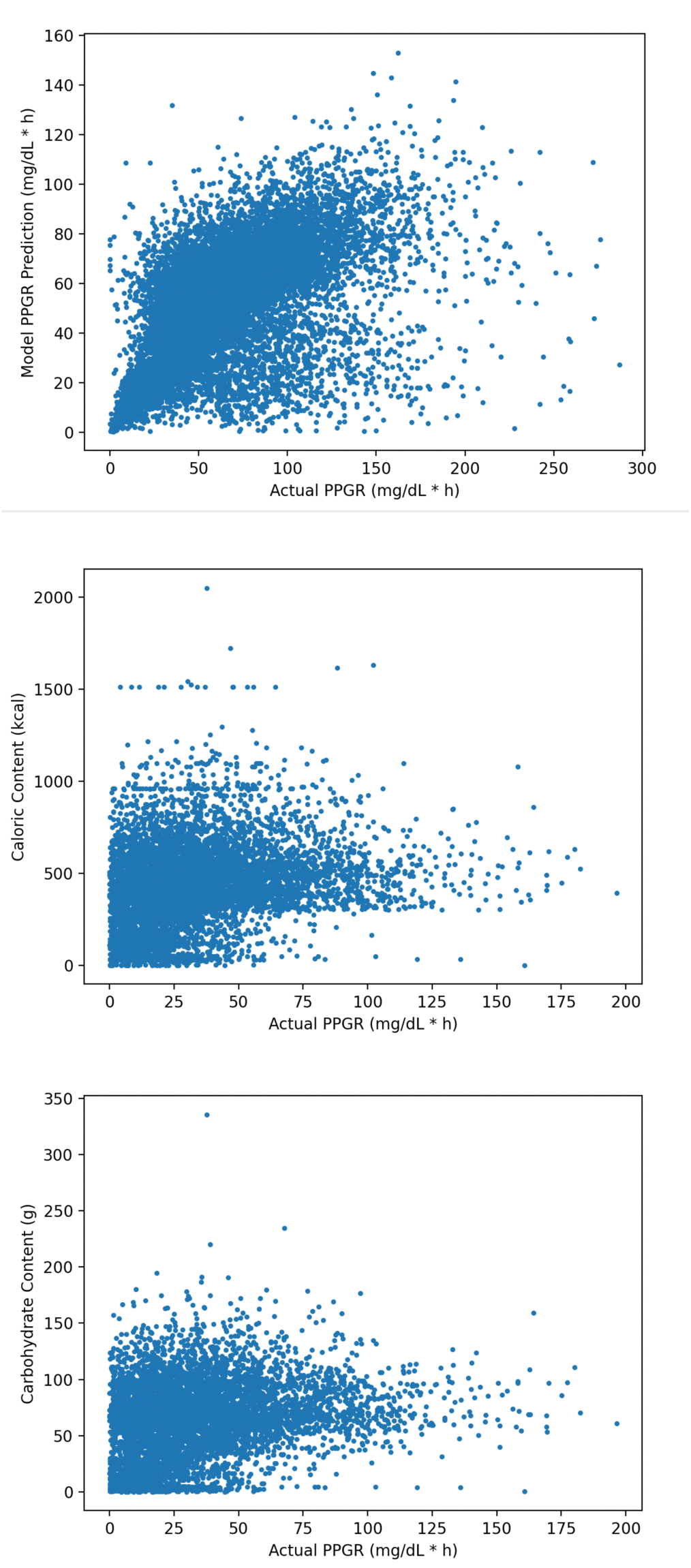
Comparison of predicted PGGR and PPGR observed from logged meals. The plots show the correlation between PPGR values predicted based on the machine learning model (top panel), calorie content (middle panel) and carbohydrate content (bottom panel).

The ability of the machine learning model to predict high PPGRs (defined as those above the 50^th^ percentile of all observed PPGRs among study participants), is shown in **Figure 5**. The machine learning model demonstrated very strong discriminative ability (AUC 0.80, p<0.001) that was substantially greater than the discriminative ability of a model based only on calorie (AUC=0.58, p<0.001) or carbohydrate (AUC 0.60, p<0.001) content.

**FIGURE 5:**
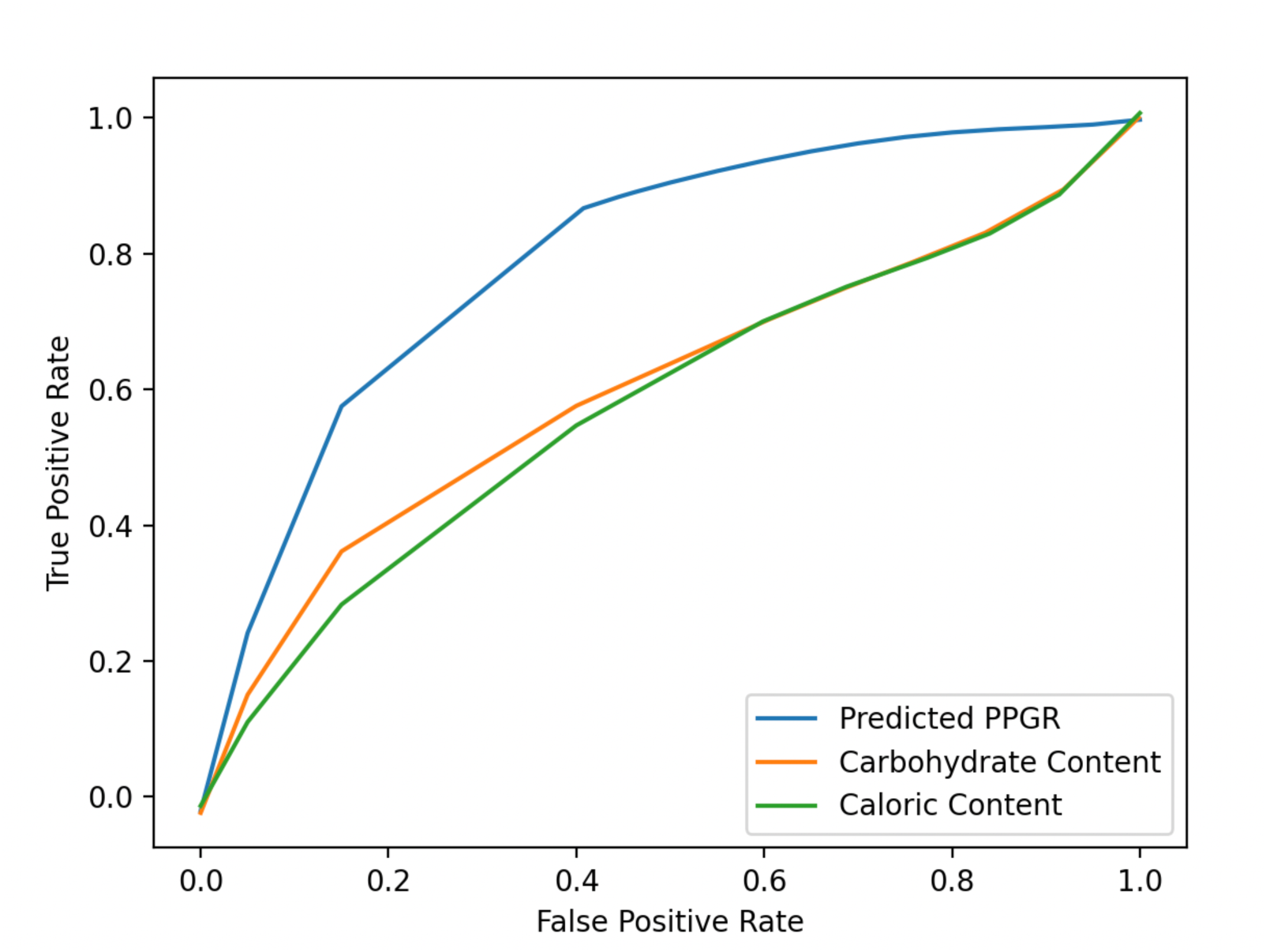
Receiver operative curve of machine learning predictions and models based on calorie and carbohydrate content only at predicting high PPGR. High PPGR was classified as the 50^th^ percentile of all measured PPGRs.

## DISCUSSION

In this study we evaluated a prospective cohort of glycemic responses to foods among Individuals with sub-optimally controlled type 2 diabetes. We observed substantial inter-individual variability to individual food components, such as carbohydrate and calorie content, as well to commonly consumed Indian foods. A machine learning model based upon nutritional content, self-reported health habits, basic biometric data and laboratory data commonly available to patients with diabetes produced highly accurate individualized predictions of PPGRs that substantially outperformed predictions based upon calorie and carbohydrate content, which are the standard approaches used in clinical practice to estimate PPGRs. These results could be used to facilitate the delivery of personalized medical nutritional therapy as is widely recommended by type 2 diabetes practice guideline in India and globally.(4–7)

Inter-individual responses to food has been previously described and attributed to differences in physical activity,(32) gut microbiome,(13; 14; 33) and genetics(34) including in variations in skeletal glucose transporters related to insulin resistance.(35) For example, a study conducted in the US among non-diabetic individuals with a mean BMI of 27 found PPGR to a standardized meal of bagel and cream cheese ranged from 6 to 94 mg/dL*h.(15) A study conducted in Israel found mean PPGR to bread and butter of 44 mg/dL*h but the bottom decile had responses of ≤ 15 mg/dL*h and the top decile has responses ≥ 79 mg/dL*h.(13) Similar data have been generated for individual without diabetes in the UK(14) and for individuals with type 1 diabetes in Israel.(36) Our results add to this literature both by studying individuals with sub-optimally controlled type 2 diabetes and by providing data from India, which has an exceptionally high burden of disease(1) and unique phenotypic features.(18–20)

Our primary goal was to develop a machine-learning model to accurately predict PPGR among individuals with type 2 diabetes. Similar models have been built in other jurisdictions although mostly among individuals without type 2 diabetes.(17). For example, a machine learning algorithm trained on CGM data, dietary, activity, anthropometrics and gut microbiota for non-diabetic individuals in Israel was much more accurate at predicting PPGR than generic models based on the carbohydrate content or the amount of calories in a meal.(13) A separate US based study had similar findings.(15) A study in the Netherlands that included a small number of individuals with type 2 diabetes along with individuals with pre-diabetes and normal glucose metabolism, a machine learning model based on CGM data was highly accurate at predicting future glucose values but this study did not specifically evaluate the ability to predict PPGR.(37) A US study of 1,000 patients of whom one-quarter had type 2 diabetes found that a machine learning model trained on CGM, HRM data and food logs was highly accurate at predicting PPGR but this study has, to our knowledge, only been published in abstract form.(17)

These studies have all relied on CGM-derived features, such as pre-meal blood sugar and the dynamics of PPGR in the post-prandial period, and other specialized data, such as fecal microbiome to make their predictions.(15) While CGMs are increasingly used, practice guidelines do not recommend their long-term use for most individuals with type 2 diabetes(38) and their acceptability and affordability remains low in lower and middle income countries such as India.(39) Accordingly, we sought to develop a PPGR prediction model based only on nutritional content, patient self-reported information, biometrics and routinely-available lab data. The resultant machine learning model had high predictive ability, both with respect to calibration and discrimination, and outperformed models based on calorie or carbohydrate content, which are the current standards used in clinical practice to estimate PPGR. This model could be used to individualize meal planning for individuals with type 2 diabetes as has been recommended by clinical practice guidelines for the management of diabetes in India and globally but which has been practically difficult to implement.(4–7)

There are several limitations to our approach. First, our design was purposely pragmatic and was intended to simulate real-world circumstances for individuals with type 2 diabetes living in India. Most notably, similar to studies conducted in other jurisdictions, we relied on self-reported dietary information. And, while we audited patient logs on an ongoing basis, there may nevertheless have been issues with protocol adherence that may have undermined the accuracy of the data we collected. Fortunately, any bias resulting from inaccurately reported food content would tend to bias predictions towards the null. Second, while we recruited subjects from around India, they were cared for in clinics predominantly treating diabetes and subjects were required to have functional English literacy as well as a cellphone capable of running study specific devices. Thus, our results may not be fully generalizable to patients who do not fulfill these criteria. Third, some of enrollment overlapped with the COVID-19 pandemic which may have influenced access to health care, dietary practices and glucose control for individuals with type 2 diabetes.

In summary, a machine learning model built with nutritional content, health habits, biometric information and commonly available laboratory data produced highly accurate individualized predictions of PPGRs that substantially outperformed predictions based upon calorie and carbohydrate content. These results could be used to facilitate the delivery of personalized medical nutritional therapy as is widely recommended by type 2 diabetes practice guideline in India and globally.

## Supporting information

Ethics Committee Approvals

## Data Availability

The datasets generated and analysed during the current study are not publicly available as consent was not provided by participants to share their data with third parties.

## Notes

### Competing Interest Statement

Dr. Choudhry and Mr. Swamy receive consulting fees and hold equity in Decipher Health. Dr. Priyadarshini is an employee of Decipher Health. Dr. Mehta is an employee and holds equity in Decipher Health.

### Clinical Trial

CTRI/2022/02/040619

### Funding Statement

This work is supported by Decipher Health, Inc.

### Author Declarations

This study was approved by the ethics committees at all institutions enrolling patients. A supplemental file with listing the site name, full name of the ethics committee/IRB and decision has been uploaded as a supplementary file.

