## Supplementary material for "PERSONALIZED GLYCEMIC RESPONSES TO FOOD AMONG INDIVIDUALS WITH TYPE 2 DIABETES IN INDIA: DEVELOPMENT OF A MACHINE LEARNING PREDICTION MODEL": Ethics Committee Approvals

**Site Ethics Committee Approval Information**

| Site | Ethics Committee/Institutional Review Board (IRB) | Address | Decision | Decision Date |
|---|---|---|---|---|
| Dia Care - Diabetes Hormone Clinic, Ahmedabad, Gujarat | Shrey Hospital Institutional Ethics Committee | 270/5/B, Near AMCO Bank, Stadium Circle, Navrangpura, Ahmedabad-380009, Gujarat | Approved | 17-Jan-2022 |
| Inamdar Multispecialty Hospital, Pune, Maharashtra | Ethics Committee Inamdar Multispecialty Hospital | CIMET's Inamdar Multispecialty Hospital. S.No.15, Fatima Nagar, Wanawadi, Pune-411040, Maharashtra | Approved | 4-March-2022 |
| Renova Neelima Hospitals, Hyderabad | Institutional Ethics Committee, Neelima Hospitals | Neelima Hospitals, Sanathnagar, Hyderabad-500018, Telangana | Approved | 4-Feb-2022 |
| Bhandari Clinic and Research Centre, Jaipur | Institutional Ethics Committee, Jaipur National University | Jaipur National University, Jaipur-Agra Bypass, Near New RTO Office, Jagatpura-302017, Jaipur | Approved | 25-Jan-2022 |
| Maharaja Agrasen Hospital, New Delhi | Institutional Ethics Committee, Maharaja Agrasen Hospital | Maharaja Agrasen Hospital, West Punjabi Bagh, New Delhi-110026, Delhi | Approved | 7-March-2022 |
| Ganesh Shankar Vidyarthi Memorial Medical College, Kanpur, Uttar Pradesh | Ethics Committee, Ganesh Shankar Vidyarthi Memorial Medical College | Room no. 125, 1st floor, GSVM Medical College, Kanpur-208002, Uttar Pradesh | Approved | 30-March-2022 |
| Mar Augustine Golden Jubilee Hospital, Ernakulam, Kerala | Institutional Ethics Committee, Mar Augustine Golden Jubilee Hospital | Mar Augustine Golden Jubilee Hospital, Mookkanoor (PO), Angamaly, Ernakulam-683577, Kerala | Approved | 8-Jan-2022 |
| Bangalore Diabetes Centre and Diagnostic Lab., Bengaluru, Karnataka | Medisys Clinisearch Ethical Review Board | Medisys Clinisearch India Pvt. Ltd., Bangalore Diabetes Centre, No.426, 4th Cross, 2nd Block, Kalyan Nagar, Bengaluru-560043, Karnataka | Approved | 18-Feb-2022 |
| Dayanand Medical College and Hospital, Ludhiana, Punjab | Drug Trials Ethics Committee, Dayanand Medical College and Hospital | Dayanand Medical College and Hospital, Tagore Nagar, Ludhiana-141001, Punjab | Approved | 12-April-2022 |
| Meenakshi Multispecialty Hospital, Chennai, Tamil Nadu | Chennai Meenakshi Multispecialty Hospital<br>Ethics Committee | Chennai Meenakshi Multispeciality Hospital Ltd., New No.70, Old No.149, Luz Church Road, Mylapore, Chennai-600004, Tamil Nadu | Approved | 11-March-2022 |
| Sparsh Hospital and Critical Care, Bhubaneshwar, Odisha | Institutional Ethics Committee, Sparsh Hospital | Sparsh Hospitals & Critical Care Pvt. Ltd., Plot No. A/407, Sahid Nagar, Bhubaneswar-751007, Odisha | Approved | 7-Feb-2022 |
| Medical College Kolkata, Kolkata, West Bengal | Institutional Ethics Committee, Medical College | Medical College Kolkata, 88, College Street, Kolkata-700073, West Bengal | Approved | 28-April-2022 |
| Downtown Hospital, Guwahati, Assam | Ethics Committee Downtown Hospital | Downtown Hospital, Sankardev Path, Dispur, GS. Road, Guwahati-781006, Assam | Approved | 25-Feb-2022 |
| CHL- Hospitals, Indore, Madhya Pradesh | Integrity Ethics Committee, CHL-Hospitals | CHL-Hospitals, Near L. I. G. Square, A. B. Road, Indore-452008, Madhya Pradesh | Approved | 3-March-2022 |
